# The Impact of the Variation of CT Scanner on the Prediction of human papillomavirus (HPV) Association of Oropharyngeal Cancer (OPC) using Radiomic models

**DOI:** 10.1101/2021.03.04.21252908

**Authors:** Reza Reiazi, Colin Arrowsmith, Mattea Welch, Farnoosh Abbas-Aghababazadeh, Christopher Eeles, Tony Tadic, Andrew J. Hope, Scott V. Bratman, Benjamin Haibe-Kains

## Abstract

Studies have shown that radiomic features are sensitive to the variability of imaging parameters (e.g., scanner model) and one of the major challenges in these studies lies in improving the robustness of quantitative features against the variations in imaging datasets from multi-center studies. Here, we assess the impact of scanner choice on the computed tomography (CT)-derived radiomic features to predict association of oropharyngeal squamous cell carcinoma with human papillomavirus (HPV). This experiment was performed on CT image datasets acquired with two different scanner manufacturers. We demonstrate strong scanner dependency by developing a machine learning model to classify HPV status from radiological images. These experiments revealed the effect of scanner manufacturers on the robustness of the radiomic features, and the extent of this dependency is reflected on the performance of HPV prediction models. The results of this study highlight the importance of implementing an appropriate approach to reduce the impact of imaging parameters on radiomic features and consequently on the machine learning models.

## INTRODUCTION

Recent advances in radiomics, the process of extracting descriptors from the radiological images by mathematical algorithms, have led to a large set of quantitative imaging features available to both research and clinical communities. A number of radiomics-driven computer models have shown promising results for personalized medicine, especially in oncological applications [1–4]. Radiomic features exhibit different levels of complexity and express properties of the lesion shape and the voxel intensity histogram, as well as the spatial arrangement of the intensity values at the voxel level (texture). They can be extracted either directly from the images or after applying different filters or transformations [5–7]. Quantitative features are usually categorized into discrete subgroups: Shape, Gray level Difference Method (GLDM), First Order Statistics (FO), Gray Level Co-occurrence Matrix (GLCM), Gray Level Size Zone Matrix (GLSZM), Gray Level Run Length Matrix (GLRLM) and Neighborhood Gray-Tone Difference Matrix (NGTDM). One of the main drawbacks of radiomic features is their potential lack of reproducibility due to variation in imaging parameters, which is assumed to be greater than the variation caused by either manual segmentation [8] or inter-observer variability [9]. This variation affects the information being extracted by radiomic features, which in turn affects classifier performance [10,11]. Consequently, conclusions regarding the performance of radiomic models must be treated with caution [12] since the results are vulnerable to image acquisition variability [13,14].

As radiomics is used to develop predictive models in biomedicine, it is important to assess the impact of domain dependency on the development of radiomic models using machine learning. A prediction task that has received broad attention in the literature is the prediction of human papillomavirus (HPV) association of oropharyngeal cancer (OPC) from radiological images[15–19]. HPV-positive OPC is now recognized as a distinct disease entity with implications for treatment and prognosis [20,21]. HPV status is currently ascertained from tumor tissue using immunohistochemistry to visualize expression of the p16 protein, or by using in situ hybridization for viral DNA. As such, standard HPV testing is invasive as it requires tissue sampling. Therefore, seeking a non-invasive yet accurate way to assess HPV status is an important research goal.

Recently, a statistical radiomics approach analyzing Computerized Tomography (CT) images has emerged as a potential non-invasive approach to predict HPV status in OPC patients [15,16,18]. In this study, we evaluated the impact of domain dependency made by the type of scanner manufacturer on this prediction. We leveraged a large image database compiled from consecutively treated OPC patients at the Princess Margaret Cancer Centre with the aim to assess the influence of scanner manufacturer on feature reproducibility and prediction of HPV status. We found that the scanner manufacturer affects the prediction of HPV status by a ML model built upon CT-derived radiomic features. Our results also indicate that robust features might reduce the overfitting in radiomic models and subsequently affect the accuracy of the prediction.

## METHODS

The schematic overview of this study is shown in Figure 1.

**Figure 1:**
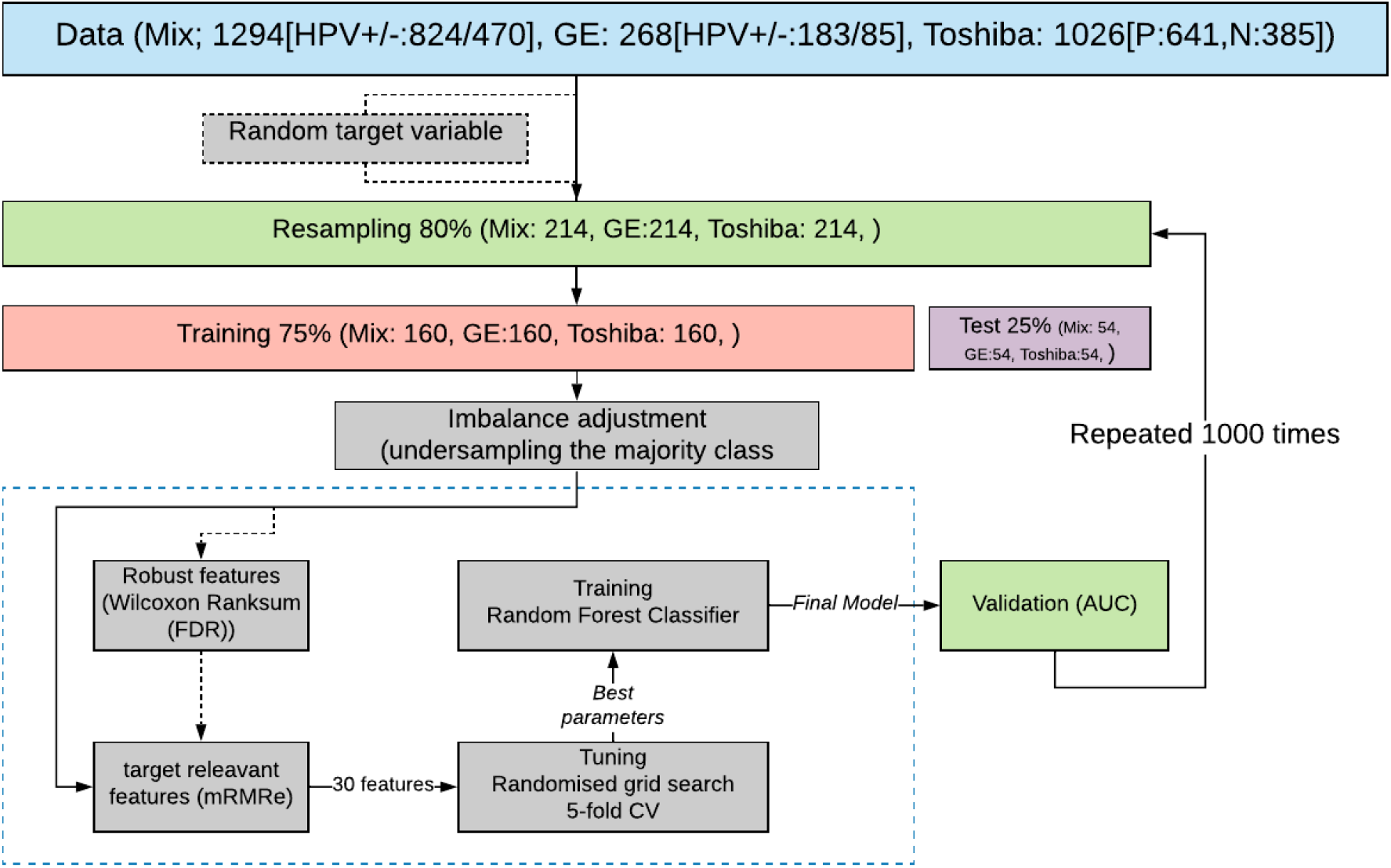
This schematic overview of this study

### Dataset

Patient data were retrospectively collected from the Princess Margaret Cancer Centre, University Health Network and was approved by the institutional review board (REB 17-5871). All experiments were performed in accordance with the relevant guidelines and regulations of the institution. The primary patient cohort in this paper was collected by searching the institutional database for consecutive in-patients who met the following criteria: (1) had Oropharynx cancer (OPC); and (2) had completed p16 immunohistochemistry. In total, we analyzed CT images from 1,294 OPC patients with known HPV status determined by p16 immunohistochemistry (Supplementary Figure 2). Mean patient age was 61.5 years ± 10.5 (standard deviation). HPV status was positive in 824 patients (641 Toshiba and 183 GE) and negative in 470 patients (385 Toshiba and 85 GE). Distribution of HPV status was almost the same in two groups (+HPV:0.78[Toshiba]/0.22[GE]; −HPV:0.81[Toshiba]/0.19[GE]). Intravenous contrast was used in 371 patients (all from Toshiba scanner). The dataset was subsequently stratified by CT scanner manufacturer (Toshiba scanner, GE scanner and both scanners (Mix)). Next, nine configurations of train-test sets were made: (1) Toshiba-Toshiba, (2) GE-GE, (3) Toshiba-GE, (4) GE-Toshiba, (5) Mix-Mix, (6) Toshiba-Mix, (7) GE-Mix, (8) Mix-Toshiba and (9) Mix-GE. Mix group contains the same number of samples from two scanner manufacturers (i.e Toshiba and GE).

### Feature extraction

For each patient, the primary gross tumor volume (GTV) was contoured by the treating oncologist. Prior to extraction, images were resampled to 1×1×1 mm voxels and the intensities were normalized with a bin width of 25. We extracted a total of 1,874 radiomic features from each patient’s manually-segmented GTV using PyRadiomics (version 3) [22]. The extracted features belong to six feature classes including: *Shape* features describing the shape and geometric properties of the region of interest (ROI) such as volume, maximum diameter along different orthogonal directions, maximum surface, tumor compactness, and sphericity. *First-order statistics* features describing the distribution of individual voxel values without concern for spatial relationships. These are histogram-based properties reporting the mean, median, maximum, minimum values of the voxel intensities on the image, as well as their skewness (asymmetry), kurtosis (flatness), uniformity, and randomness (entropy). *Second-order statistics* features including the so-called textural features [23], which are obtained by calculating the statistical inter-relationships between neighboring voxels. They provide a measure of the spatial arrangement of the voxel intensities, and hence of intra-lesion heterogeneity. Such features can be derived from the grey-level co-occurrence matrix (GLCM), quantifying the incidence of voxels with same intensities at a predetermined distance along a fixed direction, or from the Grey-level run-length matrix (GLRLM), quantifying consecutive voxels with the same intensity along fixed directions [24]. Feature breakdown according to the group they belong to is as follows: 14 Shape, 320 GLRLM and GLSZM, 360 First order, 480 GLCM, 280 GLDM and 100 NGTDM.

Features are also obtained after mathematically transforming the images through application of imaging filters with the aim of identifying repetitive or non-repetitive patterns, suppressing noise, or highlighting details. These include wavelet transform, square, square root, gradient, exponential, Laplacian transforms of Gaussian-filtered images [25]. Further explanations about the detail of the aforementioned filters can be found in PyRadiomics documentation. Distribution of features based on the imaging filter is as follows: Original (unfiltered images) 88, Exponential, Gradient, Square and Square-root each 88; Local Binary Pattern (lbp) and logarithm of Gaussian (LoG) each 264; 704 Wavelet. Finally, all the radiomic features were scaled by subtracting the median and dividing by the interquartile (the range between the 1st quartile and the 3rd quartile).

### Data sampling and splitting

Figure 1 shows the overall workflow of this study. Initially, 80% of the data was resampled without replacement and then was split into train and test sets with the proportion of 75/25. The remaining 20% was held out for final validation. Subsequently the training set has been used for feature selection (discussed later) and model training, and the resultant model has been tested on the test set. The above process has been repeated 1000 times to evaluate the statistical significance of the obtained result and the median value of the obtained performance metric has been reported (Figure 1).

### Reproducibility Analysis and feature selection

T-Distributed Stochastic Neighbor Embedding (t-SNE) clustering was applied to visualize potential scanner dependency in the radiomic features. t-SNE is a non-linear technique for dimensionality reduction that is particularly well suited for the visualization of high-dimensional datasets. The algorithm starts by calculating the probability of similarity of points in high-dimensional space and then tries to minimize the difference between these similarities for a meaningful representation of data points in lower-dimensional space. We test whether distributions of observations obtained between the two different groups on the selected variable are systematically different using the Wilcoxon rank-sum test. Our assumption was that features with the same distributions across two scanner manufacturers will have the least scanner dependency (we define these features as “robust” if they are not statistically significantly associated with scanner manufacturer). We corrected the p-values for multiple testing and computed the false discovery rate (FDR) using Bonferroni correction [26] with a threshold set at 5% for significant dependency.

### Feature selection

In order to select relevant features for HPV prediction, we used the Minimum Redundancy, Maximum Relevance (mRMR) Ensemble Feature Selection (mRMRe) implemented in the PymRMRe package (version 1.0.4) [27]. This technique is a feature selection approach that selects the features with a high correlation with the class (maximum relevance) and a low correlation between themselves (minimum redundancy). We used the F-statistic to calculate the correlation with the class (relevance) and the Pearson correlation coefficient to calculate the correlation between features (redundancy).

### Tuning and training

Imbalance adjustment was done by under-sampling the majority class (HPV positive) and Random forest classifier was trained to predict the HPV status (Figure 1). We used the GridSearchCV function in Scikit-learn (0.23.2) for exhaustive search over the specified values of the model’s hyper-parameter such as the number of trees, maximum depth of the tree, minimum number of samples required to be at a leaf node. Each model was trained on the 1000 features selected by mRMRe. Finally, RF models were trained with and without robust features. The predictive performance of the HPV status classifiers were assessed by calculating the area under the curve (AUC) (i.e., the area under the curve of receiver operating characteristics). For training, five-fold cross-validation was applied in which training sets were randomly partitioned into five groups. When one group was used for testing, then the other groups were retained for training. For each combination, the training-testing procedures were repeated 100 times until each sample in the data set was assigned a prediction score. The final AUC was estimated based on the average prediction score (1000 times). In parallel all the above processes were repeated by replacing actual target label with random binary label to compare the result with random models.

## RESULTS

In order to visualize the distribution of scanner manufacturers in high-dimensional feature space, we performed t-SNE dimensionality reduction directly on the scaled features, plus silhouette analysis for all samples. Cases have been labelled with the type of scanner manufacturer (Figure 2A). Clustering showed a dependency to the scanner manufacturer when all radiomics features were used (average Silhouette score ∼ 0.4). We also labelled the clustered data with the HPV status and found that the observed clusters were not related to the patient HPV status (Figure 2B) (average Silhouette score ∼ 0.03). We performed a Wilcoxon rank-sum test to identify features that are robust between Toshiba and GE scanners (FDR≥5%). We found that 53% (989 of 1874) of the radiomic features were significantly associated with the scanner classification (FDR<5%). We then computed the t-SNE clusters again using only the robust (FDR≥5%) features and confirmed that the data did not cluster by scanner group (Supplementary Figure 3). To illustrate the distribution of robust features, the average (over 100 separate runs) proportion of robust features according to the total number of features in each class and a total number of robust features were also estimated. Approximately, on average 740(±90) features (out of 1847) were significantly associated with the scanner manufacturer (FDR<5%). The most average number of robust features belong to the GLCM group (24±1.1%) when numbers were normalised to the total number of robust features (Figure 3A). However, when the number of robust features was normalized to the number of features in that class most of the GLDM and NGTDM (55%) features were robust against the scanner manufacturer (Figure 3C). Also for each group, the distribution of robust features after applying different image filters was compared to the original images (Supplementary Figure 4). All feature groups developed improvement in the number of robust features after applying LoG, LBP and Wavelet features implying that these filters could be of great importance in increasing features robustness. The filter groups with the largest proportion of robust features (number of robust features normalized by the total number of features in that group) were the Exponential (86%), LBP (73%) and LoG (76%) compared to original non-filter features (78%) (Figure 3B, D).

**Figure 2:**
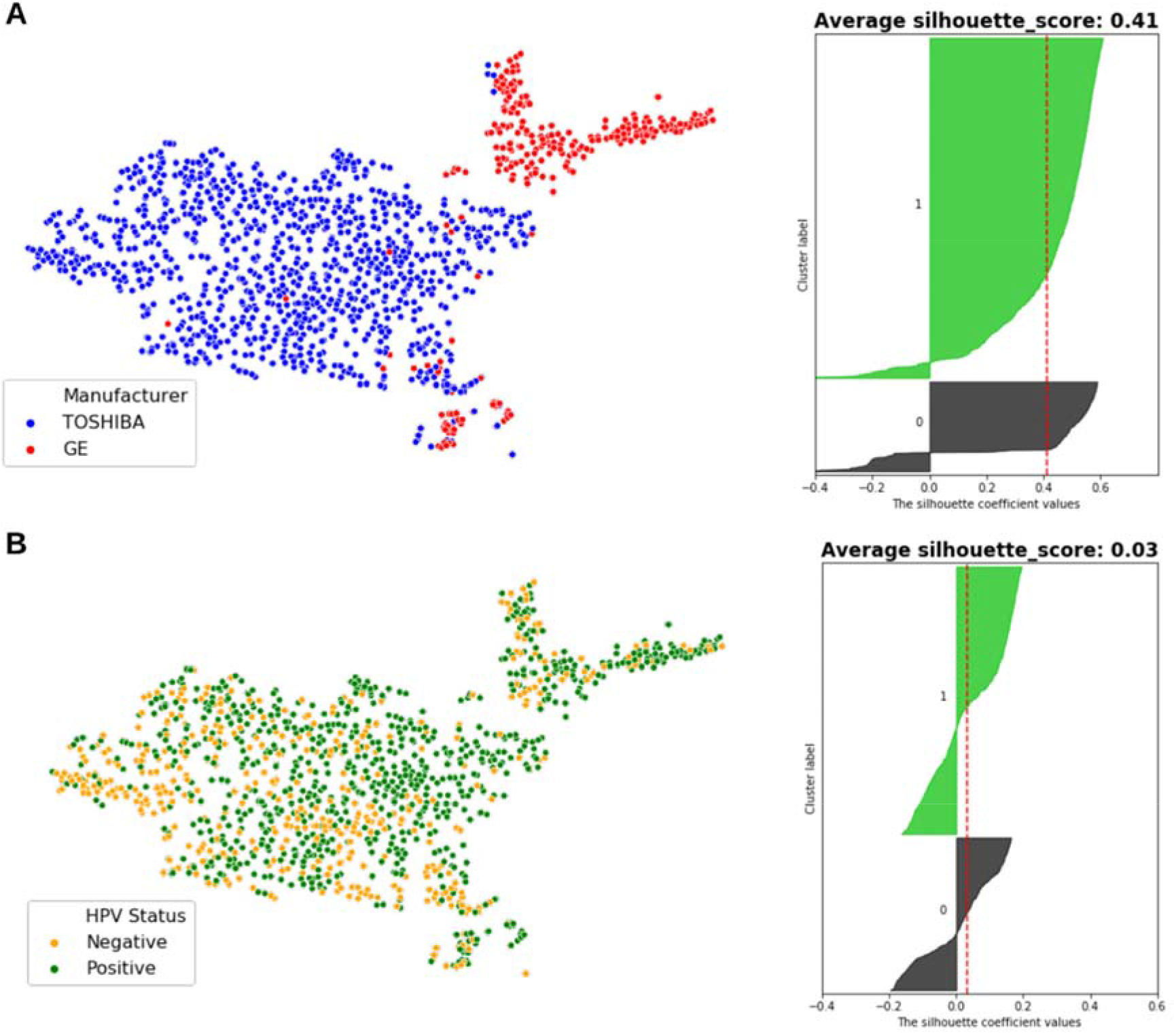
t-SNE clusters labeled by samples HPV status (A, orange: HPV negative, green: HPV positive) and scanner manufacturer (B, red: GE, blue: Toshiba). Also, the corresponding silhouette analysis and average silhouette score is shown on the right. The impact of scanner manufacturer is clearly seen when samples labeled by manufacturer type. On the hand, radiomic features do not show intrinsic dependency to the sample’s HPV status.

**Figure 3:**
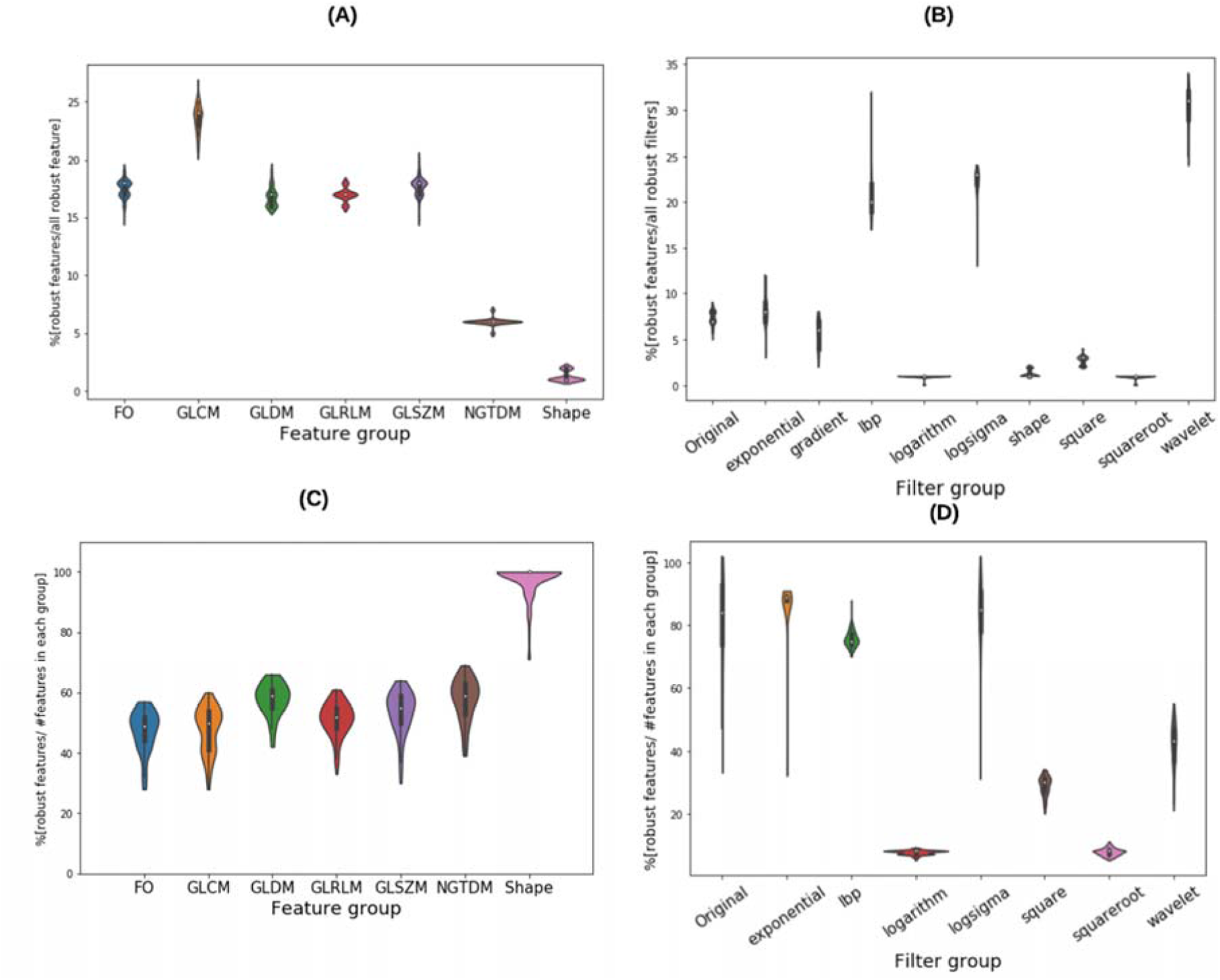
distribution of robust features according to the type of feature group (A, C) and imaging filters (D,B) normalized to the total number of robust features (A, B) and the number of features in each feature group (C,D).

The distribution of the selected robust features deemed HPV-relevant (after mRMRE feature selection) is presented based on different scanner models (GE, Toshiba and mix). The result showed in terms of feature type, first order statistics (Figure 4A) and in terms of imaging filter, Wavelet filters (Figure 4B) give rise to the largest number of robust features. The wavelet transformation is responsible for the largest number of robust features (28% of total HPV-relevant robust features). However, and after applying feature selection over the robust features, GLDM and NGTDM features comprise the largest group of HPV-relevant features (Figure 4C) emphasizing the fact that robustness study considerably changed the distribution of the selected feature selection which can subsequently be reflected on the performance of the final classifier to predict HPV status. Regarding imaging filters, selecting robust features did not influence the distribution of HPV-relevant features (Figure 4D).

**Figure 4:**
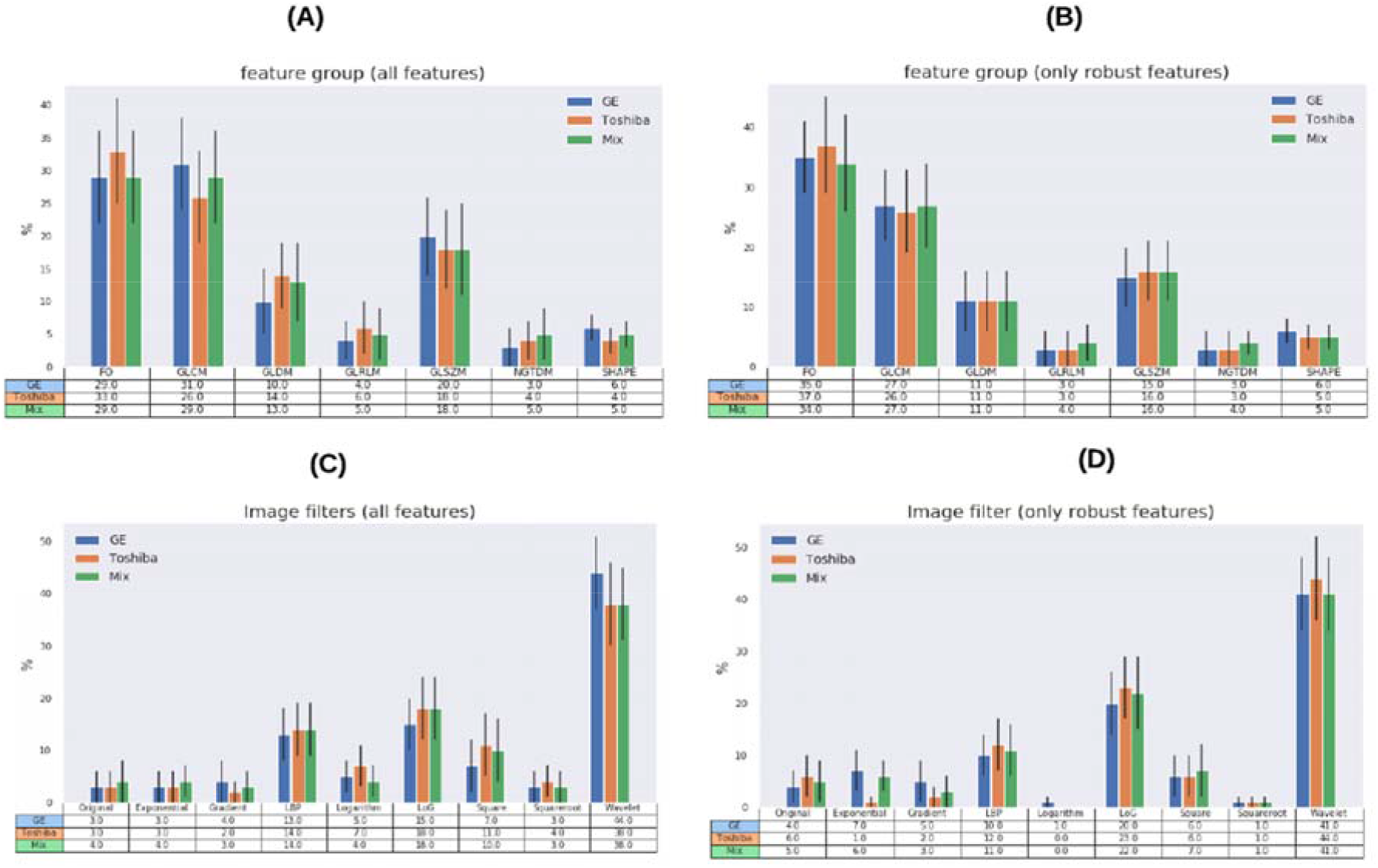
distribution (%) of HPV relevant features for different samples (GE, Toshiba and mix) according to the type of feature group and imaging filters prior to robustness evaluation (A,B) and after that (C,D).

We also evaluated the number of common features selected from different groups (i.e Toshiba, GE and Mix) out of all the available features (Figure 5A) and robust features (Figure 5B). As it was shown in the venn diagram (Figure 5), 7 (p-value <1E-3) features were found to be common across different scanners when all features were used for modelling. This number increased to 14 (p-value <1E-3) when only robust features were used. The number of common features between Toshiba-GE, Toshiba-Mix and GE-Mix are 1,16,0 when all features used for feature selection and 0,14, 2 when only robust features have been applied. As it was obvious robustness increased the number of common features among all groups.

**Figure 5:**
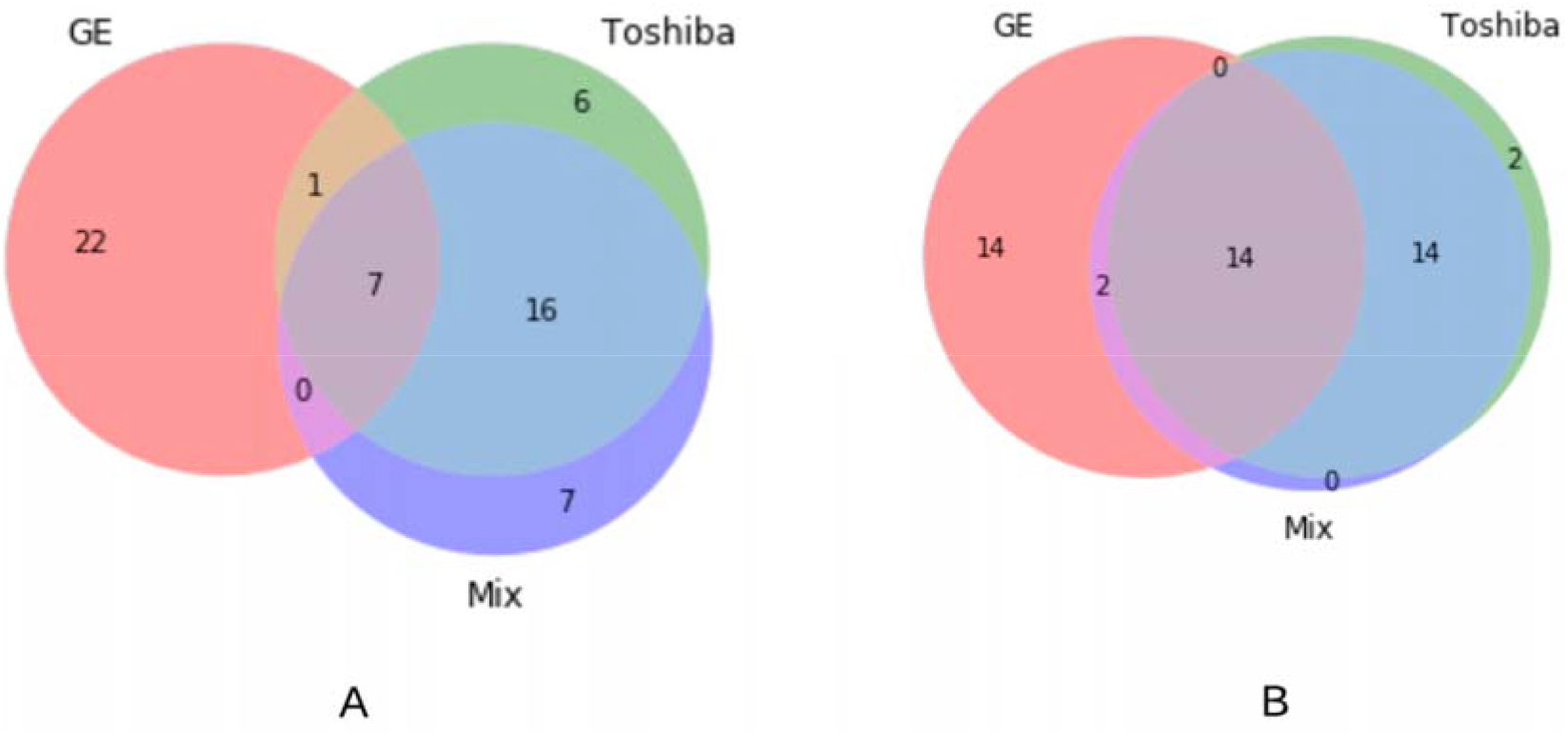
Venn diagram of the common radiomic features selected out of samples from different CT scanner type from all radiomic features (A) and only robust features (B)

### Scanner Grouping and Prediction of HPV status

The highest and lowest mean AUC values were 0.79 (p-value < 1E-4) and 0.70 (p-value: 5.4E-3) and obtained with Toshiba-Mix and Toshiba-GE respectively (Figure 6).

**Figure 6:**
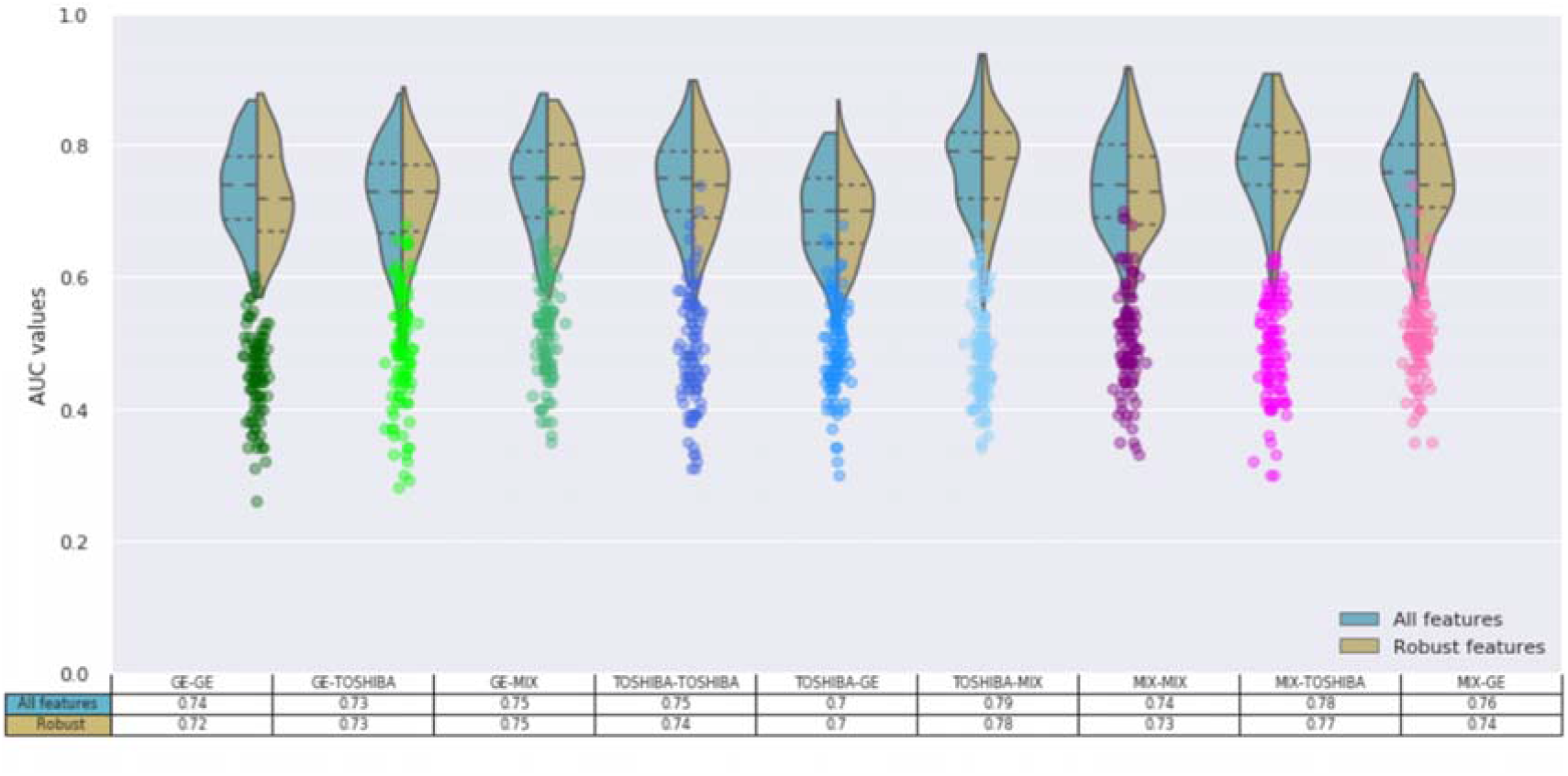
the prediction accuracy (AUC-ROC) of HPV status obtained by the Random Forest Classifiers for 9 configurations of scanner manufacturers, used for training and tests after 100 runs. The Wilcoxon rank-sum test applied to select robust features against the scanner models (adjusted p-value > 1E-2, Bonferroni correction). The mRMRe was used to select HPV relevant features. The model was trained and tested on different sets based on their scanner manufacturer (T: Toshiba, G: GE, mix) with a different number of features (mRMRe and mRMR + Robust). The corresponding scatter plots are from the same model but random dependent variable.

For models trained on one scanner manufacturer, the highest and lowest result in terms of median AUC obtained when they were tested on Mix sample (i.e GE-Mix [0.75, p-value:4E-4], Toshiba-Mix [0.79,p-value <1E-4]) and other scanner manufacturers (i.e GE-Toshiba [0.73, p-value: 7E-4], Toshiba-GE [0.70,p-value: 5.4E-3]) respectively.

The Random Forest model was trained and tested on both samples(Mix) and reached a mean training and validation AUC of 0.79(p-value <1E-4), 0.74 (p-value:4E-4) respectively. Also, this model was trained on robust features (FDR≥0.05) and reached a mean AUC of 0.77(p-value < 1E-4), 0.73(p-value: 4E-4) in training and validation respectively. This result reveals that robust features tend to reduce the difference between training and validation AUC which can be best described as reduction in models overfitting. Also models trained on one scanner manufacturer but tested on Mix samples resulted in AUC values of 0.78 (p-value <1E-4) and 0.76 (p-value: 6E-4) for Toshiba-Mix and GE-Mix models respectively. The training AUC in all models reduced after removing non-robust features (GE: 0.80→0.77, Toshiba:0.81→0.79, Mix: 0.79→0.77).

The models with single scanner manufacturers resulted in not significantly different AUC value (GE-GE:0.74 [p-value < 1E-4], Toshiba-Toshiba:0.75 [p-value:6E-4]) than both scanners (Mix-Mix: 074.). Also, after removing non-robust features, the Mix-Mix model reached a train and validation AUC of 0.77(p-value < 1E-4), 0.73 (p-value: 4E-4) (Figure 6).

## DISCUSSION

Our goal was not to find a model that led to a good classification of HPV status but to find the impact of different CT scanners on the prediction performance of the radiomic model. To do this, we assessed the effect of different scanner manufacturers on the robustness of radiomic features and their use for the prediction of HPV status in OPC, an increasingly common type of head and neck cancer. Although there are many studies investigating the robustness of radiomic features, a few reported the impact of feature robustness on the predictive performance of radiomic models. In this study, the scanner manufacturer affects the prediction accuracy of the HPV status using hand-engineered radiomics features.

Scanner dependency is an important aspect of radiomics research that has previously been evaluated in phantom studies [12,28]. In these studies, the researchers using CCR phantom images from different scanners by different manufacturers concluded that most features have significant scanner dependency and pointed out the importance of minimizing this effect in future radiomics studies. Other studies highlighted that different CT scanners have been proven to have variation in their Hounsfield units even with the same acquisition parameters [29,30]. Perrin et al. showed that when they included all the patients from all scanners, the number of liver tumor-derived robust features (concordance correlation coefficient > 0.9) from the same scanner model decreased from 75 to 35 (out of 254) [31]. This retrospective study evaluated the impact of scanner manufacturers on the prediction of HPV status using CT-derived radiomic features. To the best of our knowledge this is the first study evaluating scanner dependency using patient data.

To evaluate the effect of domain dependency in the prediction of HPV status, Random forest classifiers trained and tested on samples from different scanners (GE vs Toshiba vs. Mix). A total of 1,874 radiomic features extracted from the GTV of 1,294 OPC patients. The t-SNE clustering and the Wilcoxon rank-sum tests were then utilized to visualize the dependence of radiomic features on scanner manufacturers. This allowed for quantitatively measuring the statistical variation between features from each scanner manufacturer. The t-SNE clustering showed that radiomic features are dependent on the scanner manufacturer.

We found that most of the robust features belong to the GLCM group, which was in accordance with previous studies [32,33]. In a study to evaluate the variations of radiomic features extracted from 20 NSCLC patients from different scanners, Busyness and texture strength of the NGTDM class were the most and least robust features, respectively [12]. Based on the definition in [34], NGTDM textural features reflected the intensity differences between a voxel and its neighboring voxels. With the exception of Wavelet imaging, filters do not significantly change the distribution of robust features from the non-filtered images (Original). One main reason behind the superiority of Wavelet filters could be the superiority in the number of features (744 vs 93) which may overestimate this filter. However, Wavelet features have shown interesting applications in radiomics studies mostly because of their potential to highlight hidden texture information [35].

Finally, different combinations of samples from different scanner manufacturers (GE, Toshiba, and Mix) have been resampled to evaluate the effect of scanner manufacturer on the prediction of HPV status. We identified the best prediction model that yielded the best AUC equals to 0.79 was the Toshiba-Mix configuration along with the use of all the radiomic features for training. One interesting result of this study is that the robust features did not improve the accuracy of the models trained on one scanner manufacturer (GE or Toshiba) nor the accuracy of the models that used both scanners (Mix). A hypothesis behind this finding might be that over-fitting dominates radiomics models and incorporating only robust features removes any significance. Although this finding is highly dependent on the outcome of interest and subject to change with other endpoints, it highlights the applicability of feature harmonization techniques [36], providing they can be applied to new samples.

We also found that the bias in the results in favour of one scanner manufacturer (Toshiba). The optimal prediction accuracy was achieved when the training set included only one specific type of scanner compared to other scanners in all the same configurations. However, removing non-robust features reduced the accuracy of the prediction.

The current study has multiple limitations. First, we did not have the same patient imaged on the two groups of scanners, which is the standard approach in this type of study; as a result we were not able to use the common reproducibility metric used in other similar studies for variables such as Intraclass correlation (ICC) [8], Concordance Correlation coefficient (CCC) [9] or Coefficient of Variation (COV) [37]. However, this is acceptable since we were dealing with real patient data, and it is not currently feasible to collect this number of samples (1294 patients) with HPV status and two sets of images from different scanner manufacturers. Another limitation was the samples from this scanner (Toshiba) have undergone contrast agent administration while the other group being non-contrast examinations. Although the GTV area is a very small region, we believe that the contrast media administration is a major contributor to the clustering since it significantly affects the CT Hounsfield values and can variably change internal CT numbers within tumors by highlighting regions with more/less contrast uptake and/or vasculature. The effect of contrast enhancement has been studied in the delayed phase of CT images for NSCLC patients and it showed that radiomic features are substantially affected. Furthermore, the variability of radiomic features due to contrast uptake was found to be dependent largely on the patient characteristics [38]. However, in this study, we focused on the effect of domain dependency on the prediction performance disregarding the exact difference between the domains.

## CONCLUSION

In this study, the scanner manufacturer grouping affects the prediction accuracy of the HPV status using hand-engineered radiomics features. The optimal prediction accuracy was achieved when the training set included only one specific type of scanner (i.e. Toshiba) which reflects a bias in radiomic features towards the scanner type and/or scanning methods used on that device. This result demonstrated the importance of imaging parameters, such as hardware parameters and protocols, for training radiomic-based classifiers. Future directions of this study are to evaluate how this finding will translate into clinical applications of radiomic models and potential solutions such as feature harmonization to remove this scanner dependency.

## Data Availability

The code for all the computation generated during the study are available from the corresponding author on reasonable request.

## ACKNOWLEDGMENTS

Research reported in this study was supported by Canadian Institute of Health Research (CIHR) under grant number: **426366**

## SUPPLEMENTARY FIGURES

**Figure 1.**
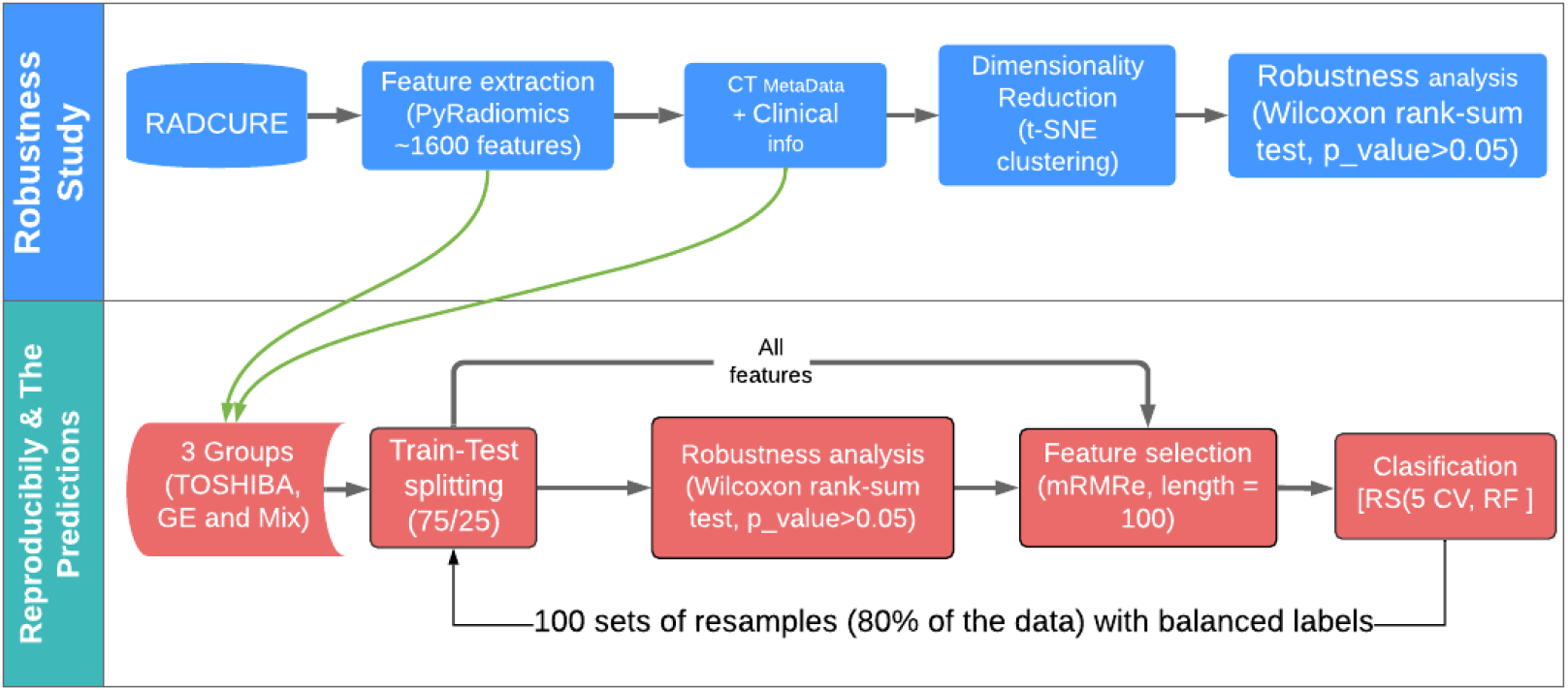
Study flowchart. Phase one of the study (Reproducibility study): First, features are extracted from CT images. Next, the robustness of features is investigated by means of Wilcoxon rank-sum test and t-SNE clustering (top row). In phase two, machine learning models were built upon different configurations of input samples to evaluate the impact of robust features on the prediction of HPV status.

**Figure 2.**
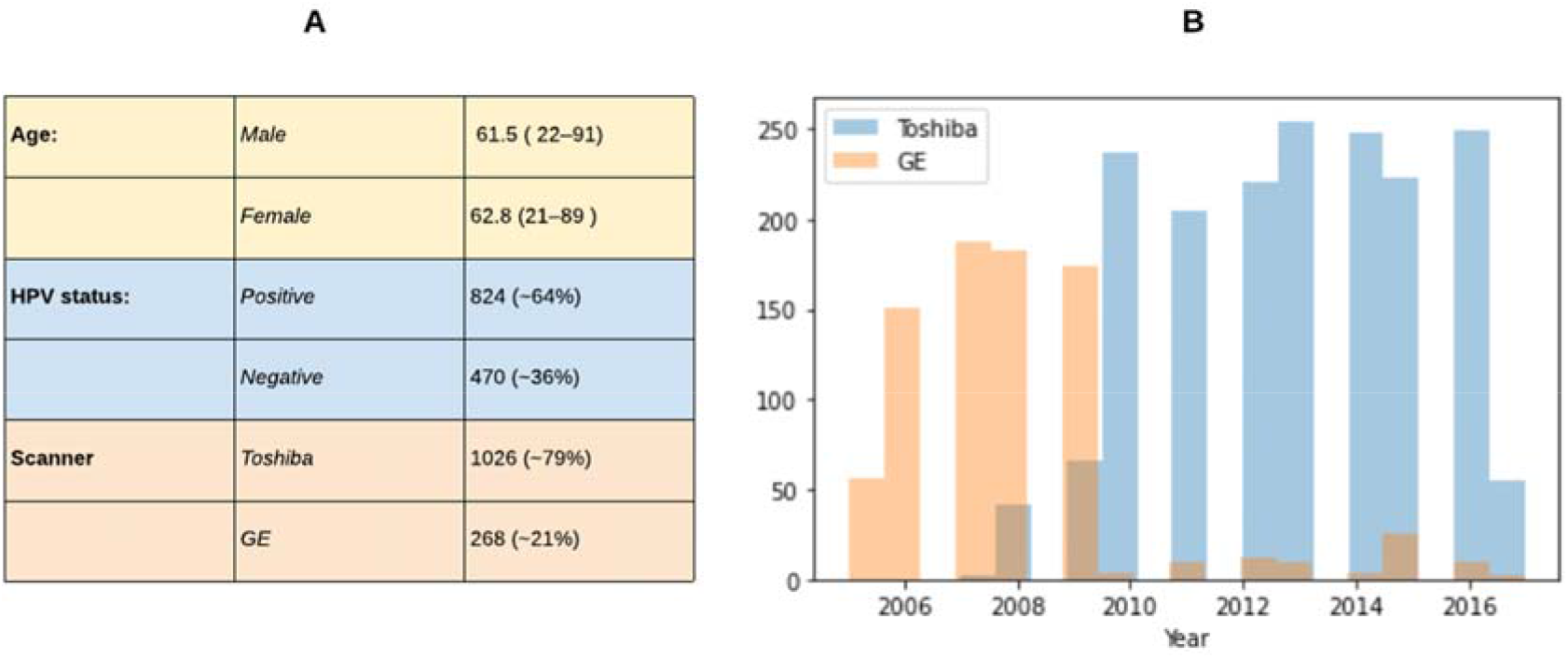
Distribution of patient base on demographyc information (A) and the type of scanner manufacturer over time (B)

**Figure 3.**
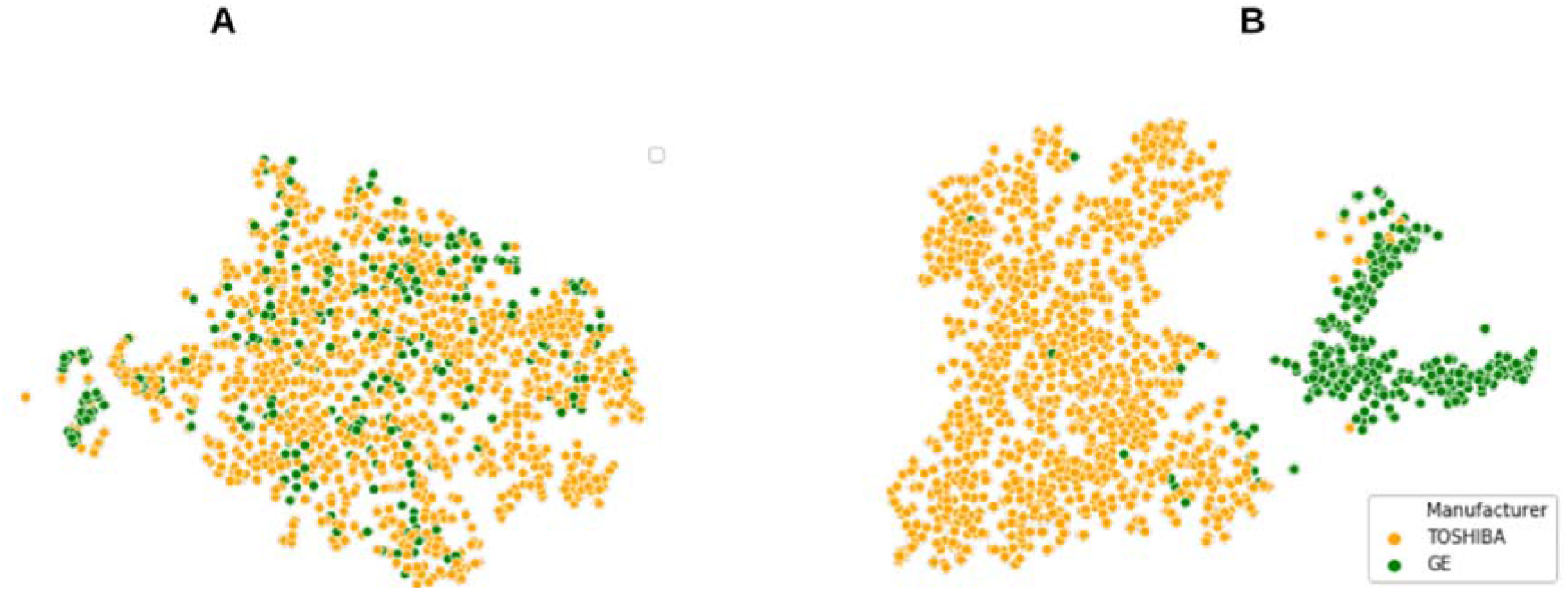
t-SNE clusters: A: robust (Wilcoxon rank-sum test, p-value > 0.05, corrected for the number of features using Bonferroni method); B: non-robust features.

**Figure 4.**
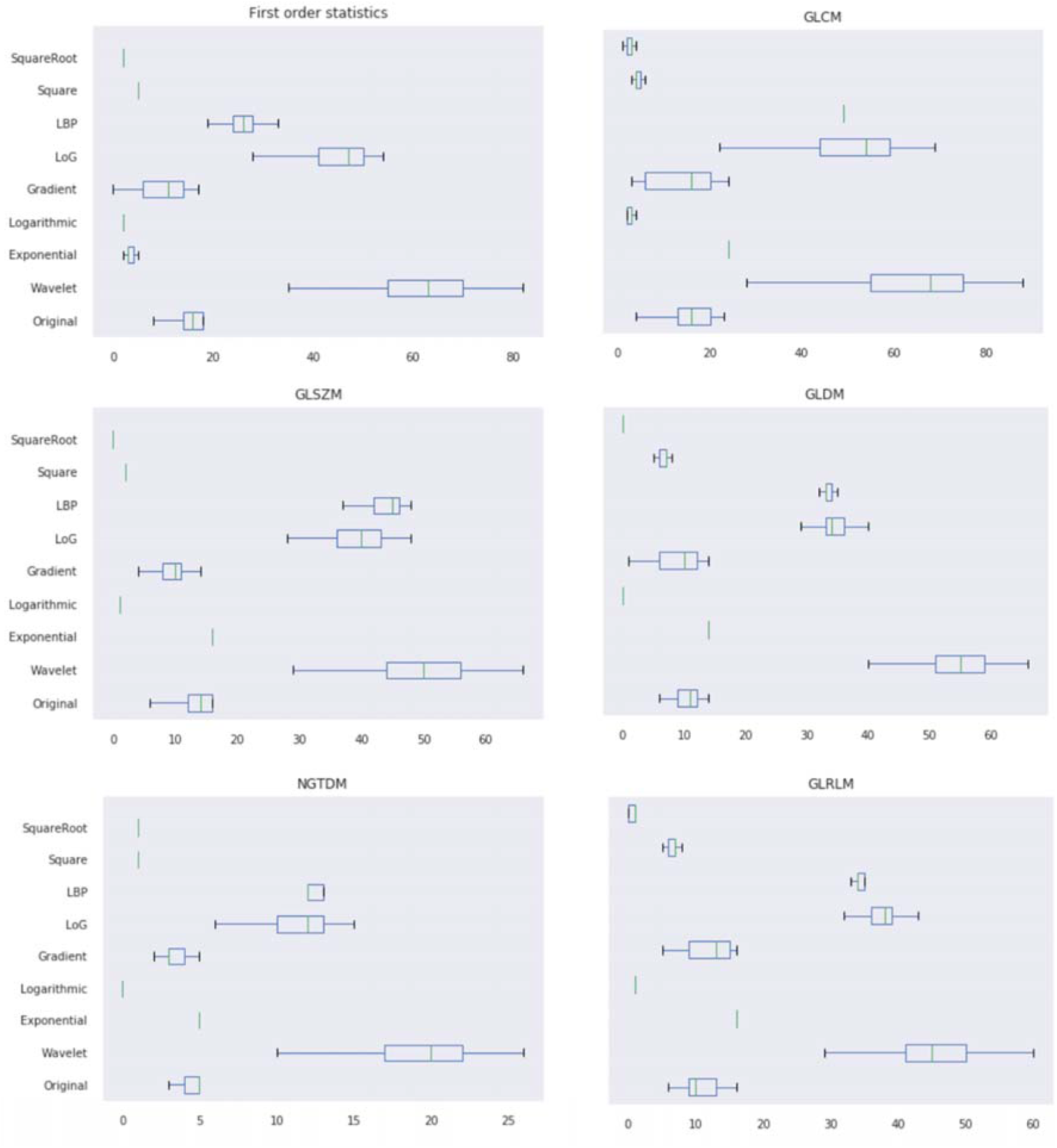
The proportion of robust features with different image filters. Values were normalized to the total number of features in each category.

**Figure 5.**
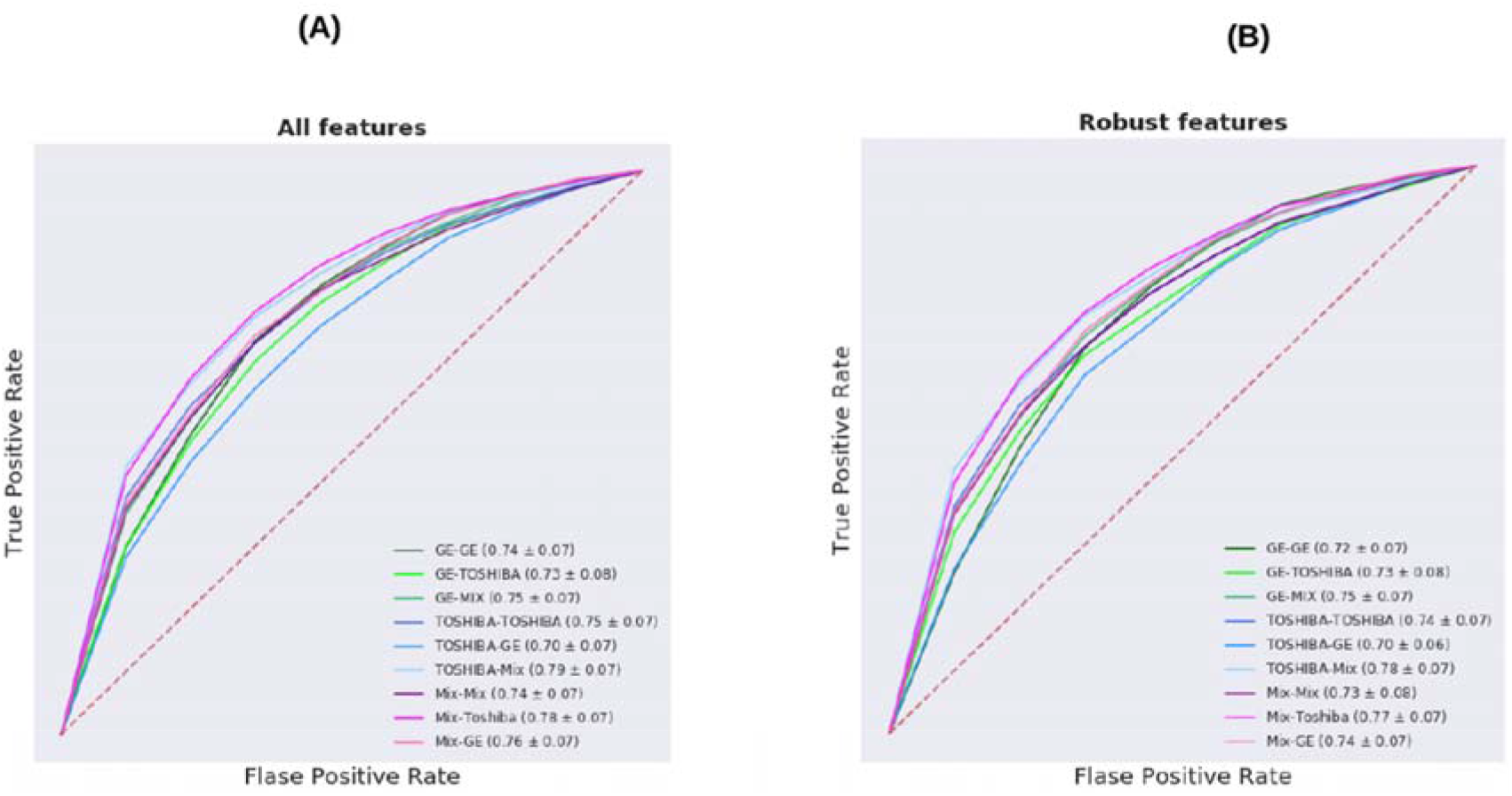
Average Receiver Operating Characteristic (ROC) curve of different models over 100 separate runs. Models are built over the HPV relevant features regardless of the robustness (A) and robust features (Wilcoxon Rank-sum test) (B). The first part of the model name stands for the type of training set and the second part represents the type of test set. The average of the AUC values was shown inside parentheses.

## Notes

### Competing Interest Statement

The authors have declared no competing interest.

### Author Declarations

Patient data were retrospectively collected from the Princess Margaret Cancer Centre, University Health Network and was approved by the institutional review board (REB 17-5871).

